# Psychological Contributors to Pain Before, During, and After Endodontic Procedures: A Scoping Review

**DOI:** 10.1101/2024.08.07.24311445

**Authors:** Atieh Sadr, Ali Gholamrezaei, Amy G. McNeilage, Cameron L. Randall, Flavia P. Kapos, Christopher C. Peck, Claire E. Ashton-James

## Abstract

**Background:** Despite an increasingly biopsychosocial approach to pain management in healthcare, the scope of research into the role of psychological factors in endodontic pain is unknown.

**Objectives:** This study aimed to identify the scope of research into psychological contributors to pain associated with endodontic procedures, as a first step towards addressing psychological contributors to pain in clinical practice.

**Method:** This scoping review was conducted and reported according to the JBI Manual for Evidence Synthesis and the Preferred Reporting Items for Systematic Reviews and Meta-Analyses (extension for scoping reviews). The data search was conducted in MEDLINE, EMBASE, PsycINFO, Web of Science, Scopus, Cochrane Database of Systematic Reviews, and CINAHL databases. For gray literature, we reviewed reference lists, medRxiv pre-prints, ProQuest and EBSCO theses, ClinicalTrials.gov and Cochrane trials (via Ovid), and conference materials via Web of Science and Scopus (from inception to July 2023). Each record was screened by two independent reviewers. Data were extracted by one reviewer and cross-verified by a second reviewer.

**Results:** Forty studies were included in the review. Twelve broad psychological constructs were evaluated in relation to pain for pre-procedural, procedural and post-procedural endodontics: pain expectancies, positive treatment expectancies, depression, anxiety, positive and negative mood (affect), beliefs about pain, desire for control of dental treatments, perceptions of dentists, somatic focus or awareness, pain coping strategies, personality, and psychiatric diagnoses. Pre-procedural pain was most frequently associated with anxiety. Procedural pain was consistently associated with expected pain. Post-procedural pain was associated with expected pain, depression, beliefs about pain, positive treatment expectations, and personality characteristics.

**Conclusion:** A variety of psychological factors have been investigated in relation to endodontic pain. Whilst associations between endodontic pain and psychological constructs were found, further research is needed to evaluate the strength of these associations, and the scope of evidence for interventions designed to address these psychological contributors to pain in dental practice.

**Registration:** The search protocol was registered on Open Science Framework in 2021 (DOI number: 10.17605/OSF.IO/FSRJP).

## 1. Introduction

Effective pain management is crucial in clinical endodontics (Cunningham & Mullaney, 1992). Research consistently links pain during dental procedures, including endodontics, to complications, persisting pain, and flare-ups (Harrison, Baumgartner, & Svec, 1983; Sathorn, Parashos, & Messer, 2008; Walton & Fouad, 1992). Current guidelines for dental pain management focus primarily on pharmacological approaches (Smith, Marshall, Selph, Barker, & Sedgley, 2017): Non-steroidal anti-inflammatory drugs (NSAIDs) are recommended in combination with acetaminophen as first-line treatments (Iranmanesh, Parirokh, Haghdoost, & Abbott, 2017; Shirvani, Shamszadeh, Eghbal, & Asgary, 2017; Smith et al., 2017), and opioids like codeine and oxycodone as second-line treatments (Iranmanesh et al., 2017; Shirvani et al., 2017; Smith et al., 2017). However, in consideration that up to 58% of patients report moderate to severe pain during endodontic procedures (Sathorn et al., 2008), and 20% report post-operative pain, there may be a need to extend our approach to pain management in dentistry beyond pharmacological treatments (Harrison et al., 1983).

Recognizing pain as a biopsychosocial phenomenon, influenced by physical, psychological, and social factors, is an important step towards improving pain management in dentistry (Raja et al., 2020). A large body of research across a range of healthcare settings demonstrates the modulation of pain by psychological factors including anxiety, depression, beliefs about pain (e.g., pain catastrophizing) and beliefs about medications (Turk & Okifuji, 2002), although there is relatively less research into social contributors to pain (c.f., Che, Cash, Fitzgerald, & Fitzgibbon, 2018; Karran, Grant, & Moseley, 2020). At present, the scope of research into the role of psychological factors in the experience of endodontic pain is not known. Although we have identified systemtatic reviews of research into anxiety in endodontic settings(Wide Boman et al. 2013, Khan et al. 2016, Chen et al. 2019), we are not aware of any research that has systematically searched for research documenting an observed relationship between psychological variables (including anxiety) and endodontic pain.

The objective of this study was to address this knowledge gap, by applying a systematic scoping review methodology, to identify and synthesise available research documenting the relationship between psychological factors (broadly defined) and pain experienced before, during, or after endodontic treatments. This review seeks to enhance our understanding of the psychological contributors to endodontic pain with a view towards identifying strategies for optimising pain management in this specialized field. A secondary aim of the present research is to assesses whether the available data on the relationship between psychological factors and endodontic pain is sufficient for meta-analysis. Due to concerns about feasibility, the current review excludes an examination of social contributors to pain.

## 2. Materials and Methods

To uphold rigorous standards, our scoping review adhered to the Joanna Briggs Institute (JBI) Reviewer’s Manual (Peters et al., 2020) and PRISMA Extension for Scoping Reviews (PRISMA-ScR) reporting guidelines (Tricco et al., 2018). The research protocol was registered on Open Science Framework (DOI number: 10.17605/OSF.IO/FSRJP).

### 2.1. Eligibility Criteria, population, concepts, and context

Based on the registered review objective, only studies examining the relationship between psychological factors and pain associated with endodontic procedures were included. Consequently, to be considered for inclusion, studies had to meet all the following criteria: 1) involving patients undergoing endodontic procedures; 2) evaluating at least one psychological factor before, during, or after the endodontic procedure; 3) evaluating pre-procedural, procedural, or post-procedural pain, and 4) evaluating the association between the psychological factor(s) and pain. We included various study designs such as cohort, case-control, cross-sectional, clinical trials (interventional, observational), systematic reviews, and meta-analyses. in English-language studies. Due to feasibility, only studies published in English were included. Case reports, case series, animal studies, letters, comments, editorials, and non-systematic reviews were not included in this review.

### 2.2. Search Strategy

Keywords were identified by reviewing relevant articles and in consultation with a librarian. We conducted a pilot search on a random sample of 50 records retrieved from PubMed, and achieved 80 percent agreement in screening (Peters et al., 2020). The search strategy was then refined based on discrepancies identified during team discussions with experts in the field (Appendix 1).

We performed a search of multiple databases, including MEDLINE (via PubMed), EMBASE (via Ovid), PsycINFO (via Ovid), Web of Science (all databases), Scopus, Cochrane Database of Systematic Reviews (via Ovid), and CINAHL (via EBSCOhost). For the Gray Literature, we searched reference lists of the included studies as well as pre-prints available on medRxiv, theses via ProQuest Dissertations and Theses/EBSCO Open Dissertations, clinical trial registers such as ClinicalTrials.gov and the Cochrane Central Register of Controlled Trials (via Ovid), and conference materials including abstracts and proceedings via the Conference Proceedings Citation Index (via Web of Science) and Scopus (from inception to July 2023). To ensure the final search strategy adhered to the recommended guidelines, we employed the Peer Review of Electronic Search Strategies (PRESS) Checklist (Tricco et al., 2018).

### 2.3. Study Selection

We used EndNote and Covidence Systematic Review software for duplicate removal and screening. Literature selection followed predetermined eligibility criteria through a two-pass screening of titles, abstracts, and full texts, resolving disagreements through consensus by two reviewer or third-reviewer intervention.

### 2.4. Data Extraction

There was no methodological quality or bias evaluation of studies included, in line with JBI (Peters et al., 2020) guidelines for scoping reviews. A single reviewer extracted the data, with a second reviewer cross-verifying data in a random sample. Key information was recorded in a charting table for a descriptive summary aligned with the aims and objectives of the review.

## 3. Results

The screened search ended with 40 articles for the final data extraction (Figure 1).

**Figure 1:**
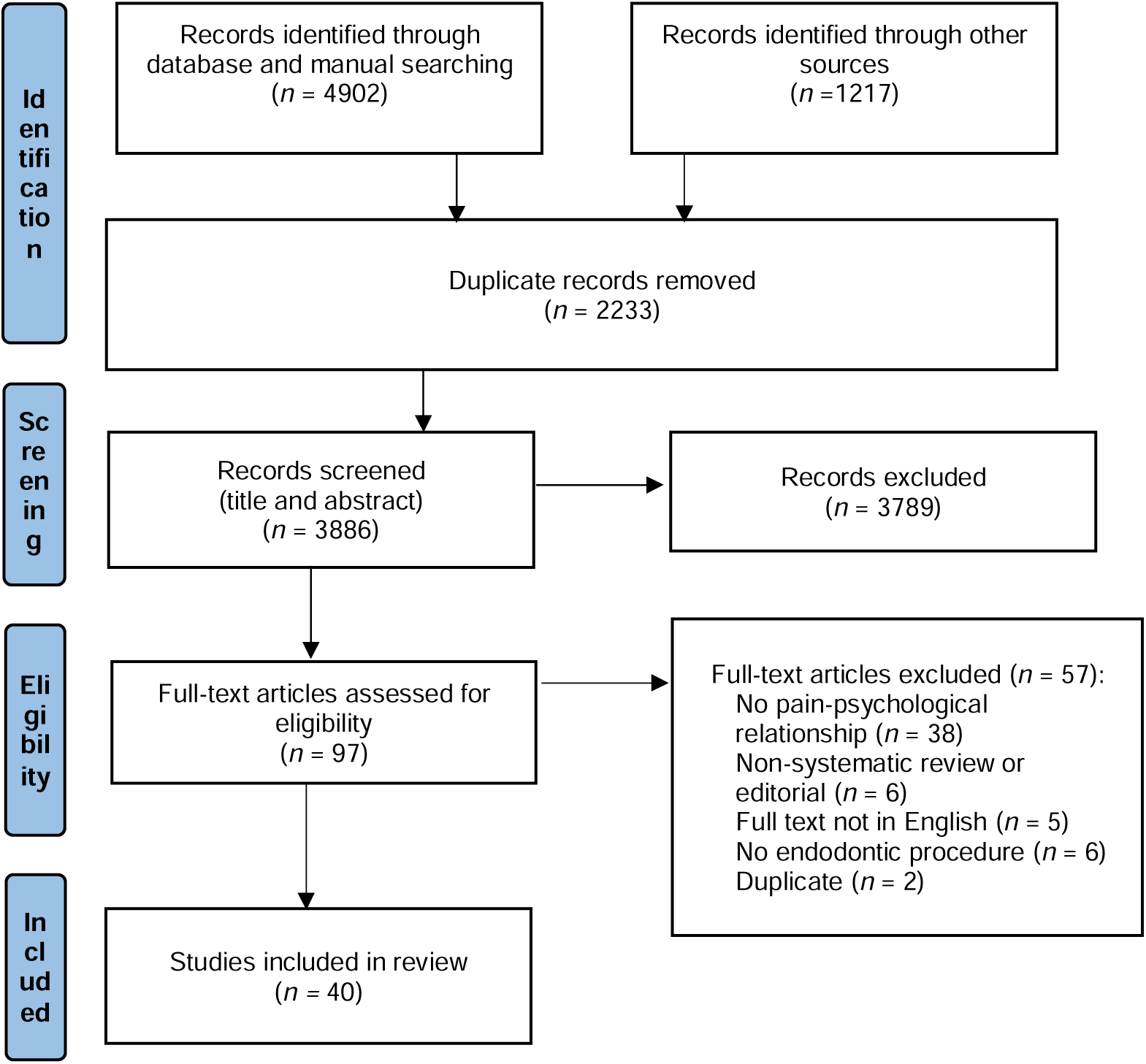
Flow chart of the studies selection process and screening

### 3.1. Characteristics of the Selected Studies

The majority of the studies (over two-thirds) were observational and prospective. The remaining studies included four retrospective studies, four cross sectional studies, and four clinical trials. The majority of studies (n=14) were from North America (the United States and Canada), with 10 conducted in Europe (United Kingdom, Germany, France, Spain, Croatia, The Netherlands, Sweden, and Denmark), 8 in Asia (Australia, China, India, Japan, Korea, Singapore, Taiwan), 4 in the Middle East (Saudi Arabia, Israel, Iran) and 4 across multiple continents (Figure 2).

### 3.2. Study Participants

Studies’ sample sizes ranged from 30 to 300 for observational studies and 1500 to 4800 for national surveys. Participants’ age ranged from 6 to 86 years (mostly adults). There were 55% female on average, with 80% of patients coming from university educational dental clinics/hospitals and the remaining 20% from private surgeries.

### 3.3. Pain Measurement

Various self-reporting scales measured pain at different stages of the endodontic procedures (pre-procedural, procedural, and post-procedural). Pain intensity, frequently assessed by the Visual Analog Scale (VAS), was the most common outcome measured (Figure 3).

### 3.4. Psychological Constructs

Data extracted from 40 studies are summerised in Table 1. Studies investigated psychological variables associated with pre-procedural pain (n=8), procedural pain (n=18), and post-procedural pain (n=20). We organised these psychological variables under 12 higher-level psychological constructs, including “anticipated or expected pain”, “positive treatment expectation”, “depression”, “anxiety” (encompassing state or trait anxiety, dental anxiety, fear, pain-related anxiety [i.e., pain catastrophising, fear of pain, psychological disability, social disability, and psychological discomfort], oral health-related anxiety; social functioning, and stress), “beliefs about pain management” (encompassing belief about the link between stress and pain and expectations about pain medications), “desire for control of dental treatments”, “perceptions of dentists” (including empathy), “somatic focus or awareness”, “positive and negative affect” (often described as mood), “pain coping strategies”, “personality”, and “psychiatric diagnosis”. Appendix 2 describes the scales used by the included studies to measure the psychological variables.

**Table 1:**
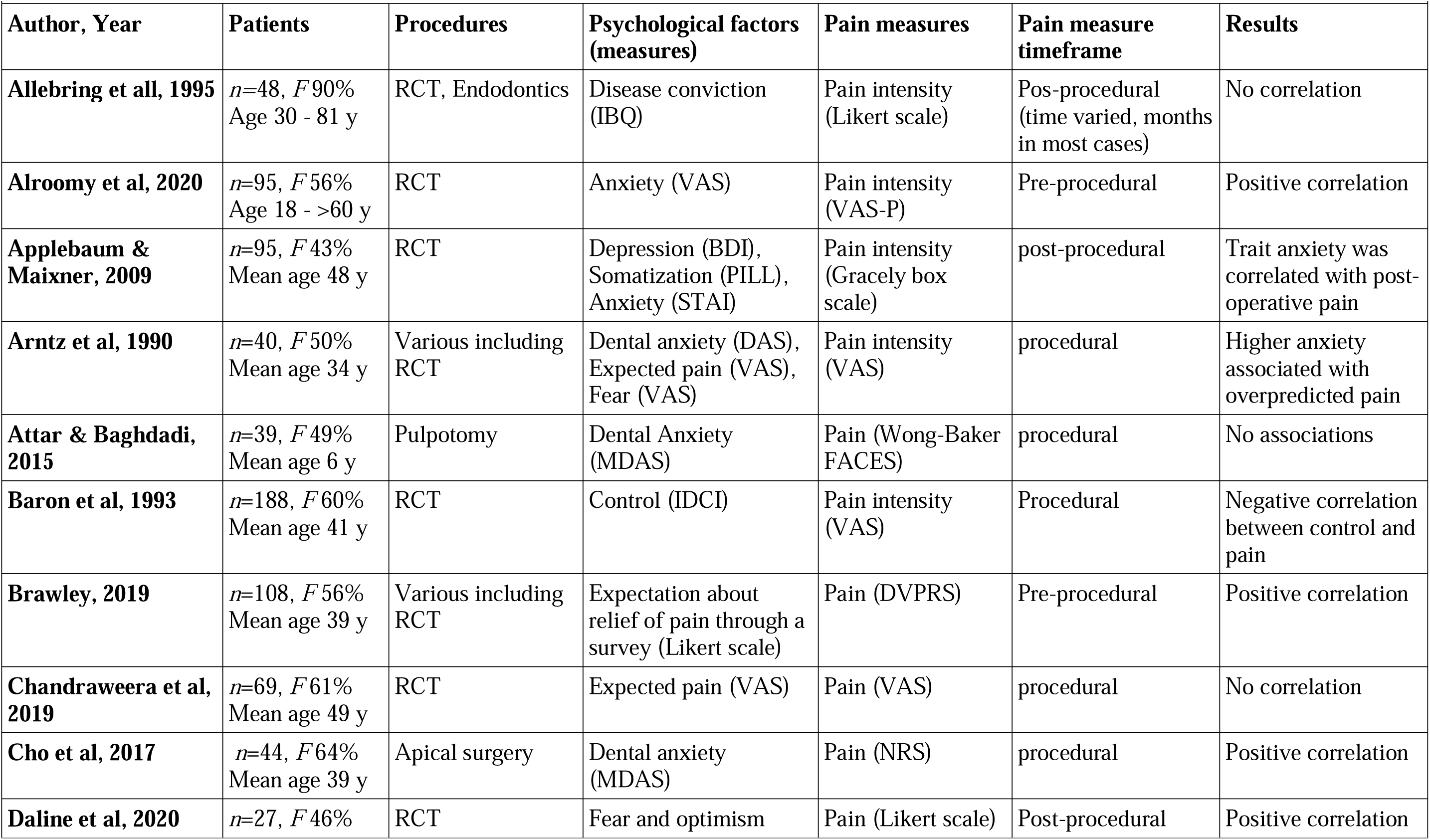
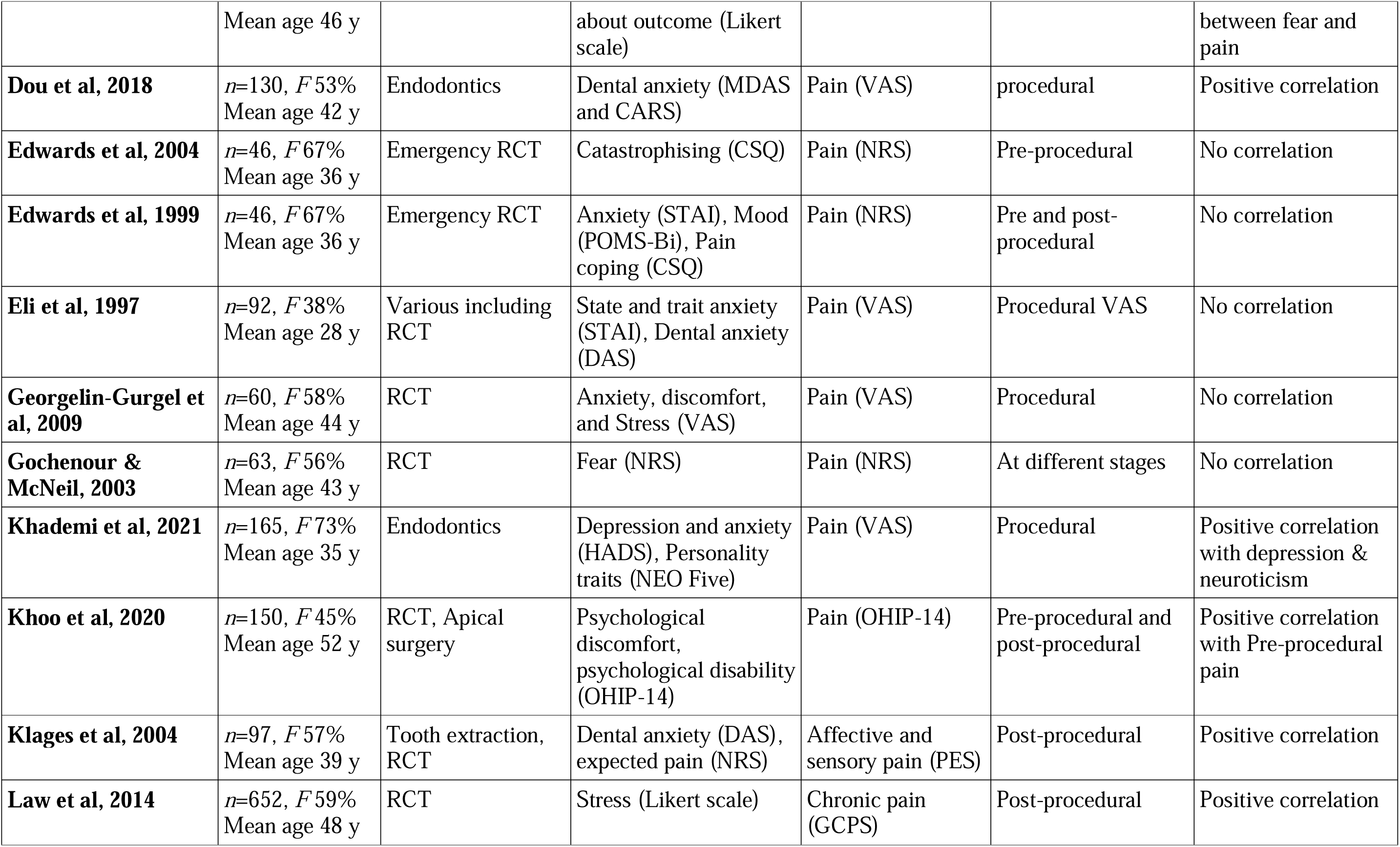
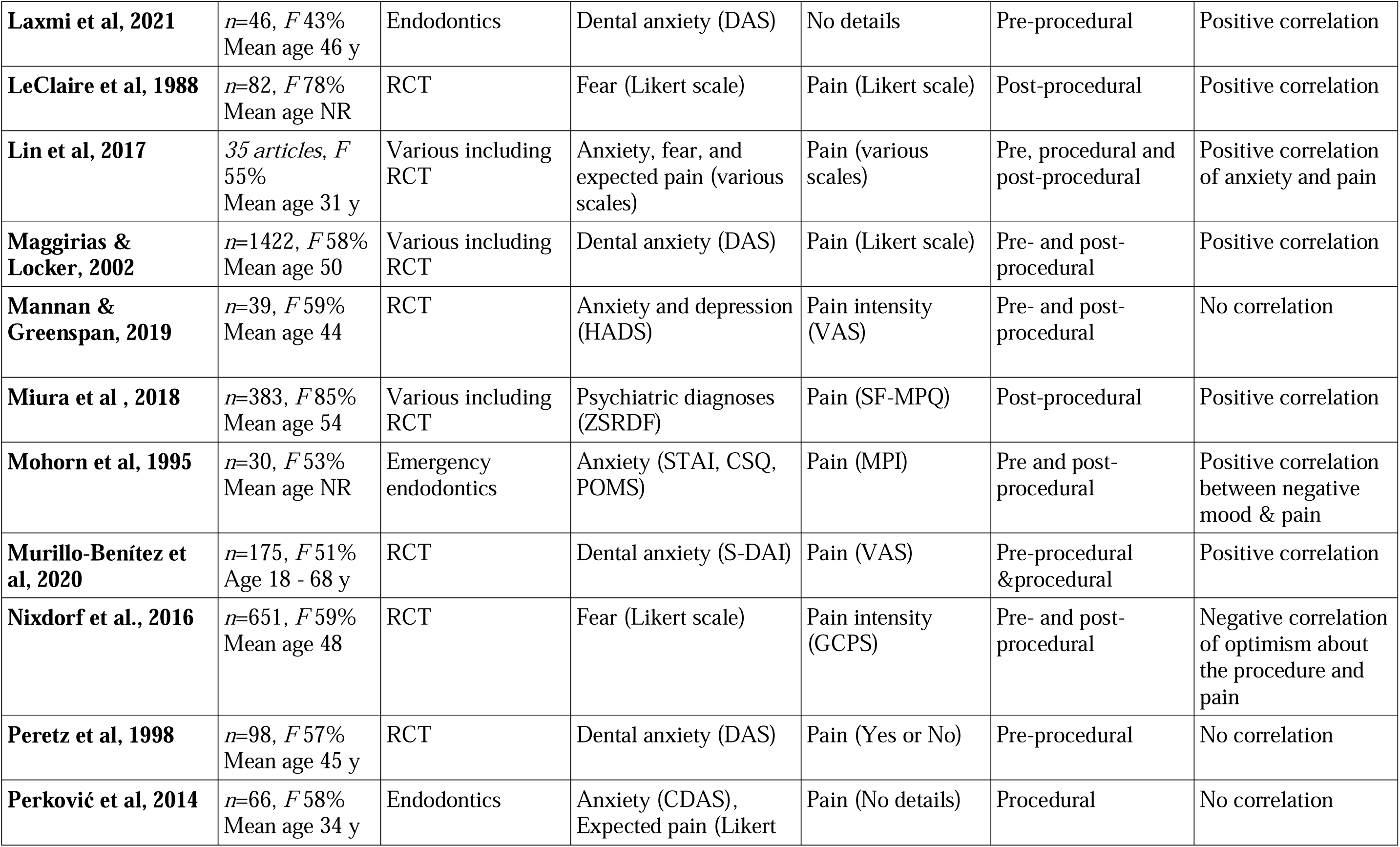
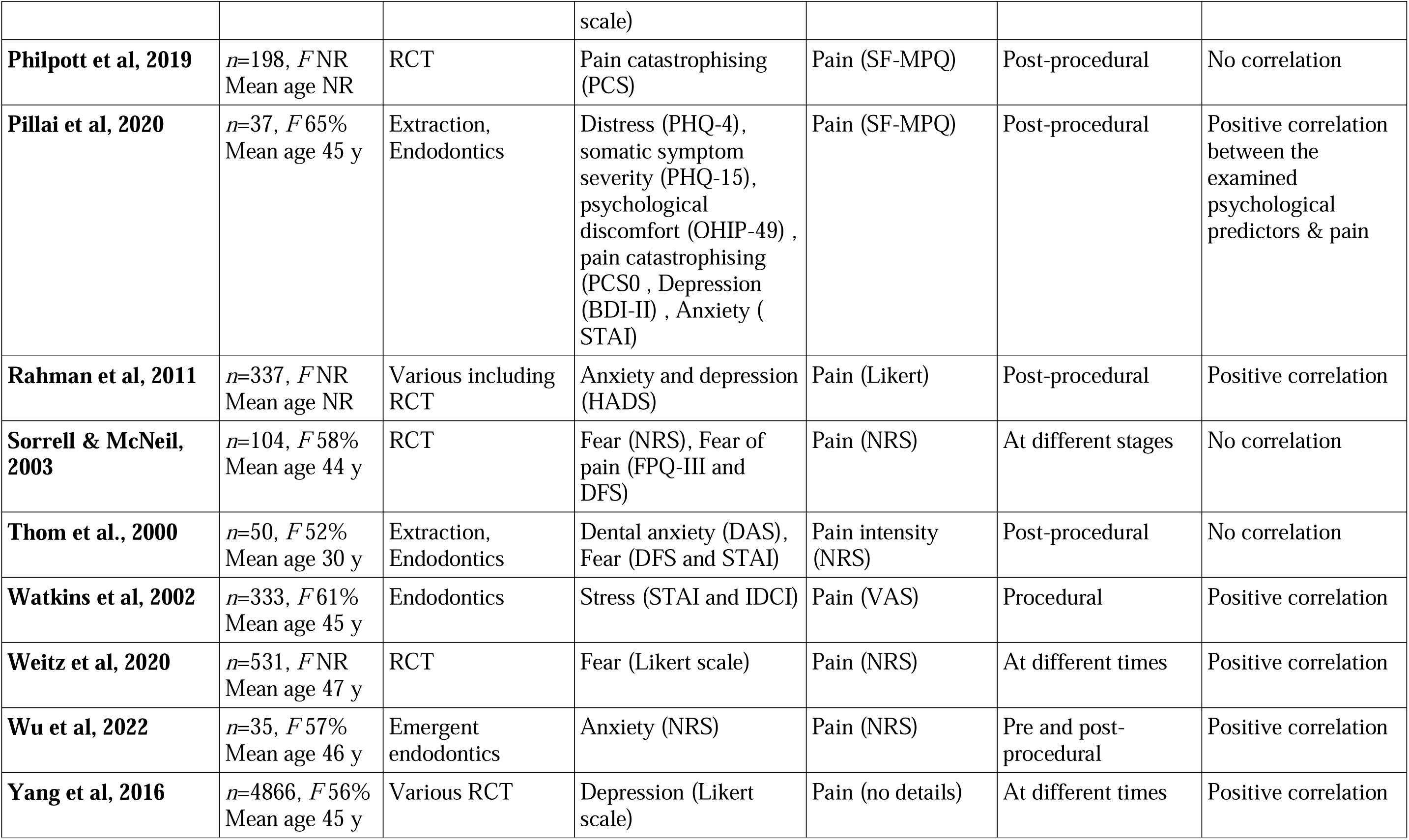
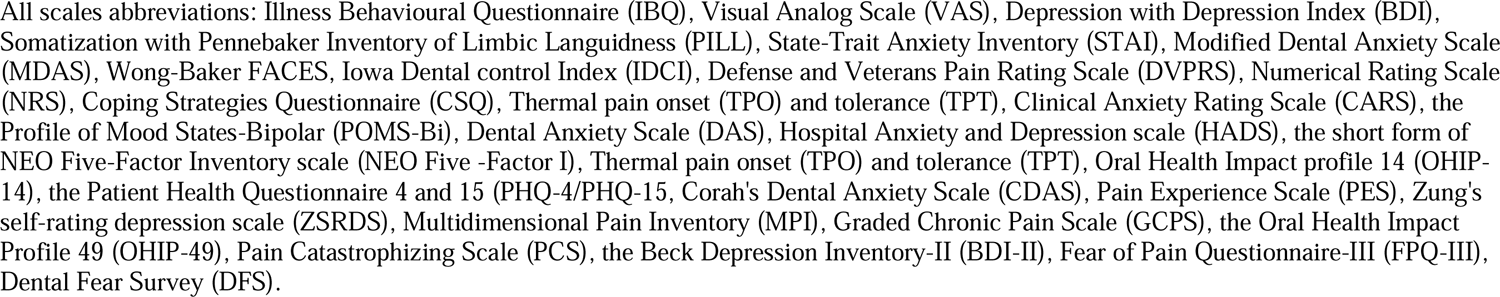
Details of Data Extraction.

### 3.5. Psychological Constructs Associated with Pre-procedural Pain

Two psychological constructs - anxiety and beliefs about pain - were investigated in association with pre-procedural pain (Alroomy et al., 2020; Brawley, 2019; Khoo et al., 2020; Laxmi et al., 2021; Murillo-Benítez et al., 2020; Peretz & Moshonov, 1998; Sorrell, 2003; Wu et al., 2022).

#### 3.5.1. Anxiety

Seven studies investigated various forms of anxiety in relation to pre-procedural pain. State anxiety was measured in two studies (Alroomy et al., 2020; Wu et al., 2022), while dental anxiety was measured in five studies (Laxmi et al., 2021; Murillo-Benítez et al., 2020; Peretz & Moshonov, 1998; Sorrell, 2003). One study (Khoo et al., 2020) measured oral health-related anxiety. Five reported a correlation between anxiety and pre-procedural pain (Alroomy et al., 2020; Khoo et al., 2020; Laxmi et al., 2021; Murillo-Benítez et al., 2020; Wu et al., 2022).

#### 3.5.2. Beliefs about Pain Management

One study (Brawley, 2019) measured patient perceptions of pain relief and expectations of receiving an opioid medication together with pre-procedural pain and reported a positive relationship between the expectation of receiving an opioid medication and pre-procedural pain.

### 3.6. Psychological constructs associated with procedural pain

Most studies (n=18) investigated the association between psychological variables and procedural pain (Arntz et al., 1990; Attar & Baghdadi, 2015; Chandraweera et al., 2019; Cho et al., 2017; Dou et al., 2018; Eli et al., 1997; Georgelin-Gurgel et al., 2009; Gochenour, 2003; Khademi et al., 2021; Klages et al., 2004; LeClaire et al., 1988; Mannan, 2019; Murillo-Benítez et al., 2020; Perković et al., 2014; Sorrell, 2003; Thom et al., 2000; Watkins et al., 2002; Weitz, 2020). The five psychological constructs investigated in these studies were anxiety, depression, expected pain, positive treatment expectations, and personality.

#### 3.6.1. Anxiety

Sixteen studies investigated the association between anxiety and procedural pain (Arntz et al., 1990; Attar & Baghdadi, 2015; Chandraweera et al., 2019; Cho et al., 2017; Dou et al., 2018; Eli et al., 1997; Georgelin-Gurgel et al., 2009; Gochenour, 2003; Khademi et al., 2021; Klages et al., 2004; LeClaire et al., 1988; Murillo-Benítez et al., 2020; Perković et al., 2014; Sorrell, 2003; Thom et al., 2000; Weitz, 2020) with mixed results. Of these, seven reported a significant positive association between anxiety and procedural pain.

#### 3.6.2. Depression

Two studies (Khademi et al., 2021; Mannan, 2019) investigated and found a positive correlation between deppresion and procedural pain.

#### 3.6.3. Expected pain

Expected pain was evaluated in five studies (Chandraweera et al., 2019; Eli et al., 1997; Klages et al., 2004; Perković et al., 2014; Watkins et al., 2002) which four found a positive association between pain expectation and procedural pain.

#### 3.6.4. Positive treatment expectations

Weitz et al. (2020) was the only study in this category that measured the expected outcome of root canal therapy, reporting a significant association between higher outcome expectancy and anaesthesia failure or procedural pain.

#### 3.6.5. Personality

Personality was investigated by one study (Khademi et al., 2021), which reported a positive correlation between neuroticism scores and pain experience during needle insertion for injection or procedural pain.

### 3.7. Psychological Constructs associated with Post-Procedural Pain

Twenty studies investigated the relationship between 11 psychological Most and post-procedural pain. The psychological variables included anxiety, depression, ‘expected pain’, ‘positive treatment expectations’, ‘positive and negative effect’, ‘pain coping strategy’, ‘desire for control’, ‘perception of the dentist’, ‘somatic focus and awareness’, ‘psychiatric disease’, and ‘beliefs about pain (Applebaum, 2009; Baron et al., 1993; Daline et al., 2020; Edwards et al., 2004; Edwards et al., 1999; Mohorn et al., 1995; Nixdorf et al., 2016; Philpott et al., 2019; Pillai et al., 2020; Rahman et al., 2011; Sorrell, 2003; Thom et al., 2000; Wu et al., 2022; Yang et al., 2016).

#### 3.7.1. Anxiety

Anxiety was investigated in relation to post-procedural pain in 14 studies (Applebaum, 2009; Daline et al., 2020; Edwards et al., 2004; Khoo et al., 2020; Klages et al., 2004; Maggirias & Locker, 2002; Nixdorf et al., 2016; Philpott et al., 2019; Pillai et al., 2020; Rahman et al., 2011; Sorrell, 2003; Thom et al., 2000; Wu et al., 2022; Yang et al., 2016), with 14 different anxiety measures used. Most studies reported a significant association between anxiety and post-procedural pain.

Three studies investigated state and trait anxiety (Applebaum, 2009; Rahman et al., 2011; Wu et al., 2022). Six studies measured dental anxiety (Daline et al., 2020; Klages et al., 2004; Maggirias & Locker, 2002; Nixdorf et al., 2016; Sorrell, 2003; Thom et al., 2000) with three investigated pain catastrophising (Edwards et al., 2004; Philpott et al., 2019; Pillai et al., 2020). Oral health-related anxiety, psychological discomfort, psychological disability, and social disability investigated in this category(Khoo et al., 2020), added with discomfort, somatic symptom, and psychological disability (Pillai et al., 2020). One study measured stress (Yang et al., 2016).

Both state and trait anxiety were significantly correlated with post-procedural endodontic pain (Rahman et al., 2011; Wu et al., 2022), with trait anxiety identified as a significant predictor of pain in the first five days after endodontic treatment (Rahman et al., 2011). Four out of six dental anxiety studies showed a positive correlation between post-procedural pain and anxiety, while two found no relationship (Nixdorf et al., 2016; Thom et al., 2000). One study (Sorrell, 2003) showed a significant correlation between pain and the Dental Fear Survey-Physiological Score (DFS). Except for Pillpott et al. (2019), two out of three studies investigating pain catastrophizing reported a significant correlation with post-procedural endodontic pain.

#### 3.7.2. Depression

Six studies investigated and four reported a significant association between depression and post-procedural pain (Miura et al., 2018; Philpott et al., 2019; Rahman et al., 2011; Mannan, 2019; Pillai et al., 2020; Yang et al., 2016). Mannan and Greenspan (2019) and Rahman et al. (2011) found depression increases post-procedural pain. Pillai et al. (2020) and Yang et al.( 2016) reported higher pain in their depressed groups.

#### 3.7.3. Expected pain

Only one study (Wu et al., 2022) examined the correlation between expected pain relief and post-procedural pain finding no signficant association.

#### 3.7.4. Positive treatment expectations

Two studies examined positive treatment expectations in relation to post-procedural pain (Daline et al., 2020; Wu et al., 2022). Daline et al. (2020) reported no association between long-term persistent pain and patients’ optimism about the treatment outcome. However, Wu et al. (2022) found a positive correlation between expected pain relief and post-procedural pain reduction.

#### 3.7.5. The Desire for Control over Dental Treatment

The desire for control over dental treatment was investigated in relation to post-procedural pain in two studies (Baron et al., 1993; Maggirias & Locker, 2002), both of which reported a significant correlation between this psychological construct and post-procedural pain.

Baron et al. (1993) reported that high desire for control or felt control associated with lower pain and higher sensory focus. In addition, Maggirias and Locker (2002) reported that pain was significantly higher in participants with higher Felt Control.

#### 3.7.6. Perceptions of Dentists

Positive perceptions of the dentist was evaluated in relation to post-procedural pain in two studies (Maggirias & Locker, 2002; Perković et al., 2014) which both reported negative correlations with pain. Maggirias and Locker (2002) assessed patients’ perceptions of the dentist and how dental care was delivered during endodontic treatments. They found a negative correlation between the high reported empathy of dentists and post-procedural pain one week after treatment.

#### 3.7.7. Somatic Focus or Awareness

Three studies investigated the relationship between somatic focus or awareness and post-procedural pain with mixed results (Applebaum, 2009; Pillai et al., 2020; Yang et al., 2016). Pillai et al. (Pillai et al., 2020) measured somatic symptom severity and reported that patients with painful post-traumatic trigeminal neuropathy (PTTN) had higher scores for somatic symptoms than healthy controls. Conversely, Applebaum and Maixner (2009) measured somatization and found no significant association between post-treatment somatization and post-procedural pain.

#### 3.7.8. Psychiatric disease

One study (Yang et al., 2016) used a national survey to measure melancholy, consultation with a psychiatrist, and suicidal thoughts before various dental procedures, including root canal therapy, and found significant association between these factors and self-reported dental pain (Yang et al., 2016).

#### 3.7.9. Pain Coping Strategies

Two studies (Edwards et al., 1999; Mohorn et al., 1995) investigated pain coping with both reported significant association with post-procedural pain (Edwards et al., 1999; Mohorn et al., 1995).

#### 3.7.10. Beliefs about Pain Management

Beliefs about pain were evaluated in only one study with the results showed that the belief that pain is made worse by stress was a significant predictor of severe post-procedural pain (Law et al., 2014).

#### 3.7.11. Positive and Negative Affect

The relationship between ‘positive and negative affect’ and post-procedural pain was examined in two studies (Edwards et al., 1999; Mohorn et al., 1995). Both studies used the Profile of Mood States (POMS) questionnaire prior to emergency endodontic treatment and reported no significant association between mood and post-procedural pain.

## 4. Discussion

This review synthesises the results of forty studies investigating twelve psychological constructs in relation to pre-procedural, procedural, and post-procedural pain.

### 4.1. Study Participants

The majority of studies were conducted in high-income countries, and over 80% of participants were recruited from university educational dental clinics/hospitals and public health centres. This concentration in specific settings and regions may limit the generalizability of the findings to broader, more diverse populations.

### 4.2. Psychological contributors to pain

Anxiety, with a focus on dental anxiety, emerged as a frequently studied psychological construct, showing a significant association with patients’ pain experiences. However, it is notable that some studies found no correlation between anxiety and pain (Perković et al., 2014; Wu et al., 2022).

This review also underscores the significance of anticipated pain as an important psychological predictor for procedural and post-procedural pain. Perković et al. (2014) showed an association between anxiety and expectations of intraoperative endodontic pain. The relationship between expected and actual pain experiences has also been documented in prior studies on oral surgery (Gómez-de Diego, Cutando-Soriano, Montero-Martín, Prados-Frutos, & López-Valverde, 2014; Treister, Eaton, Trudeau, Elder, & Katz, 2017). What patients expect before treatment can affect their experience, and this holds for different stages of treatment. Additionally, these expectations can be influenced by factors such as thoughts and emotions and the patient’s memory of past experiences. For instance, anticipating pain has been connected to feeling anxious about the treatment (Gómez-de Diego et al., 2014; Treister et al., 2017), and remembering the pain felt during past procedures is related to both anxiety and what was expected beforehand (Gómez-de Diego et al., 2014; loilova & Porreca, 2014).

Most of the reviewed studies investigated association between a psycholigical construct and post-procedural pain. Anxiety emerged as the most commonly studied predictor with most studies reporting a positive correlation. This aligns with findings from a separate study investigating anxiety in other dental procedures, demonstrating a correlation between state anxiety, anticipated pain, and experienced pain (Feinmann et al., 1987). Similar positive correlations have been observed in the context of post-procedural pain in oral surgery (Navratilova & Porreca, 2014).

Depression, another impotant psychological construct in dentistry settings, was found to be associated with both procedural and post-procedural endodontic pain in most of the reviewed studies. The results of studies in other fields, such as oral surgery and implant procedures, have been mixed (Dadgostar et al., 2017; Feinmann et al., 1987).

For the 12 constructs described, the construct of personality received limited attention with only one study (Khademi et al., 2021) addressing it. Khademi’s study (Khademi et al., 2021), investigating the positive correlation of neurotic personality with post-procedural pain, yielded similar results to an oral surgery study (Feinmann et al., 1987). Consequently, further research in this area is warranted.

### 4.3. Measurement tools

Regarding psychological and pain measurement tools, inconsistencies in defining and measuring scales were evident in the reviewed studies which may complicate pooling results and meta-analysis. Moreover, documenting the timing of pain assessments is imperative for transparent and comparable research outcomes. Many of the studies reviewed assessed procedural pain after treatment, yet the exact timing was often unclear. The effects of local anesthesia may dissipate at the time of procedural pain measurement. Thoughtful study design, considering multiple time points for pain measurement and consistent assessment protocols, is vital to enhance the accuracy and comprehensibility of research findings.

### 4.3. Limitations

Limitations to this study include restricting our search to studies published in English only, and also to psychological and not social factors due to feasibility concerns. The social factors associated with pain are likely important and will require future investigations.

### 4.4. Future directions

Based on the findings of this scoping review, future research should focus on several key areas to further understand the relationship between psychological factors and pain in endodontic procedures. Comprehensive systematic reviews and meta-analyses should be conducted to quantify the association and generalize findings across diverse populations. Additionally, quality assessments of existing studies are needed to identify methodological strengths and weaknesses, aiding in the development of standardized research guidelines. It will also be important to conduct experimental, hypothesis-driven research to investigate the mechanisms by which psychological factors contribute to endodontic pain. Finally, informed by systematic reviews and experimental research, we envisage that future research will be well placed to design and test interventions to mitigate the impact of psychological factors on endodontic pain. There have been several clinical trials of interventions to address psychological factors influencing dental pain management in paediatric patients(Yan et al., 2023)(Ram et al., 2010). While the literature offers limited insight into interventions for anxiety reduction in adult dental settings, certain strategies, including ‘tell-show-do’ (Allen, Stanley, & McPherson, 1990; Berggren, Hakeberg, & Carlsson, 2000; Wide Boman, Carlsson, Westin, & Hakeberg, 2013), hypnosis (Glaesmer, Geupel, & Haak, 2015), auditory distraction through background music (Moola, Pearson, & Hagger, 2011), and cognitive therapy (Berggren et al., 2000), have shown some promise in the field of general dentistry. We envisage that future research will test these anxiety-reduction strategies on pain in endodontic settings.

### 4.5. Conclusion

Various psychological factors have been investigated and found related to pain experienced before, during, and after endodontics procedures. We have classified these psychological factors into 12 overarching constructs, among which anxiety and expected pain have emerged as the most commonly studied factors. More research is required to understand the strength of the relationship between psychological factors and pain related to endodontic procedures, and to develop strategies to manage these psychological contributors to endodontic pain in practice.

## Data Availability

All data produced in the present work are contained in the manuscript

---

**Figure 2:** Geographical distribution of studies

**Figure 3:** Details of scales used for pain measurement based on numbers

