## Appendix 1 for "Psychological Contributors to Pain Before, During, and After Endodontic Procedures: A Scoping Review"

**Appendix 1: Databases search codes**

PubMed

(pain OR pains OR painful* OR headache* OR cephalalg* OR cephalg* OR cephalodynia* OR glossodynia* OR glossalg* OR glossopyrosis OR toothache* OR tooth-ache* OR odontalg* OR neuralg* OR neurodynia OR earache* OR ear-ache* OR otalg* OR hypersens* OR hyperalg* OR allodyn* OR "tooth sensitivity" OR "teeth sensitivity" OR "dental sensitivity" OR "dentin sensitivity" OR ache OR aches OR aching OR soreness OR sore OR flare-up* OR flareup* OR Pain[Mesh] OR Pain Measurement[Mesh] OR Hyperalgesia[Mesh] OR Dentin Sensitivity[Mesh] OR Neuralgia[Mesh] OR Symptom Flare Up[Mesh]) AND (psych* OR emotion* OR emotive OR cognition* OR cognitive* OR behavior* OR behaviour* OR mental* OR motivation* OR expectanc* OR expectation* OR anticipat* OR memory OR memories OR attitude* OR belief* OR appraisal* OR uncertaint* OR predictability OR controllability OR helplessness OR self-efficacy OR perceived-control OR felt-control OR "locus of control" OR "loss of control" OR "positive experience*" OR "negative experience*" OR threat* OR feeling* OR mood* OR fear* OR anxiet* OR anxious* OR hypervigilant* OR depression OR depressive* OR guilt* OR shame* OR embarrassment* OR distress* OR stress* OR worry OR worrisome OR disgust* OR "negative affect*" OR "positive affect*" OR "affective state*" OR phobi* OR odontophobia OR panic OR concern OR concerns OR avoidance OR reward* OR reinforc* OR vicarious* OR coping* OR learning* OR adaptation* OR optimism OR pessimism OR somatization OR somatisation OR somatoform OR neuroticism OR catastrophiz* OR catastrophis* OR nervous* OR personalit* OR mindful* OR Psychiatry and Psychology Category[Mesh] OR Health Belief Model[Mesh]) AND (Endodontic* OR (("root canal*") AND (therapy OR procedure OR treatment OR retreatment OR management OR surgery OR filling OR debridement OR devitalization OR obturation OR preparation OR resection OR hemisection)) OR pulpectomy OR pulpotomy OR (pulp AND (revascularization OR regeneration OR capping)) OR "apical surgery" OR apicoectomy OR ((peri-radicular OR periradicular) AND surgery) OR Apexification OR Apexogenesis OR Endodontics[Mesh])

OVID (EMBASE)

(pain OR pains OR painful* OR headache* OR cephalalg* OR cephalg* OR cephalodynia* OR glossodynia* OR glossalg* OR glossopyrosis OR toothache* OR tooth-ache* OR odontalg* OR neuralg* OR neurodynia OR earache* OR ear-ache* OR otalg* OR hypersens* OR hyperalg* OR allodyn* OR tooth sensitivity OR teeth sensitivity OR dental sensitivity OR dentin sensitivity OR ache OR aches OR aching OR soreness OR sore OR flare-up* OR flareup* OR exp Pain/ OR exp Pain Measurement/ OR exp Hyperesthesia/ OR exp Dentin Sensitivity/ OR exp Neuralgia/) AND (emotion* OR emotive OR cognition* OR cognitive* OR behavior* OR behaviour* OR mental* OR motivation* OR expectanc* OR expectation* OR anticipat* OR memory OR memories OR attitude* OR belief* OR appraisal* OR uncertaint* OR predictability OR controllability OR helplessness OR self-efficacy OR perceived control OR felt control OR locus of control OR loss of control OR positive experience* OR negative experience* OR threat* OR feeling* OR mood* OR fear* OR anxiet* OR anxious* OR hypervigilant* OR depression OR depressive* OR guilt* OR shame* OR embarrassment* OR distress* OR stress* OR worry OR worrisome OR disgust* OR negative affect* OR positive affect* OR affective state* OR phobi* OR odontophobia OR panic OR concern OR concerns OR avoidance OR reward* OR reinforc* OR vicarious* OR coping* OR learning* OR adaptation* OR optimism OR pessimism OR somatization OR somatisation OR somatoform OR neuroticism OR catastrophiz* OR catastrophis* OR nervous* OR personalit* OR mindful* OR exp psychiatry/ OR exp behavioral science/ OR exp behavior/) AND (Endodontic* OR (root canal* AND (therapy OR procedure OR treatment OR retreatment OR management OR surgery OR filling OR debridement OR devitalization OR obturation OR preparation OR resection OR hemisection)) OR pulpectomy OR pulpotomy OR (pulp AND (revascularization OR regeneration OR capping)) OR apical surgery OR apicoectomy OR ((peri-radicular OR periradicular) AND surgery) OR Apexification OR Apexogenesis OR exp Endodontics/)

OVID (PsycINFO)

(pain OR pains OR painful* OR headache* OR cephalalg* OR cephalg* OR cephalodynia* OR glossodynia* OR glossalg* OR glossopyrosis OR toothache* OR tooth-ache* OR odontalg* OR neuralg* OR neurodynia OR earache* OR ear-ache* OR otalg* OR hypersens* OR hyperalg* OR allodyn* OR tooth sensitivity OR teeth sensitivity OR dental sensitivity OR dentin sensitivity OR ache OR aches OR aching OR soreness OR sore OR flare-up* OR flareup* OR exp Pain/ OR exp Pain Measurement/ OR exp Hyperalgesia/ OR exp Hyperesthesia/ OR exp Hypesthesia/ OR exp Paresthesia/) AND (psych* OR emotion* OR emotive OR cognition* OR cognitive* OR behavior* OR behaviour* OR mental* OR motivation* OR expectanc* OR expectation* OR anticipat* OR memory OR memories OR attitude* OR belief* OR appraisal* OR uncertaint* OR predictability OR controllability OR helplessness OR self-efficacy OR perceived control OR felt control OR locus of control OR loss of control OR positive experience* OR negative experience* OR threat* OR feeling* OR mood* OR fear* OR anxiet* OR anxious* OR hypervigilant* OR depression OR depressive* OR guilt* OR shame* OR embarrassment* OR distress* OR stress* OR worry OR worrisome OR disgust* OR negative affect* OR positive affect* OR affective state* OR phobi* OR odontophobia OR panic OR concern OR concerns OR avoidance OR reward* OR reinforc* OR vicarious* OR coping* OR learning* OR adaptation* OR optimism OR pessimism OR somatization OR somatisation OR somatoform OR neuroticism OR catastrophiz* OR catastrophis* OR nervous* OR personalit* OR mindful* OR exp Behavioral Sciences/) AND (Endodontic* OR (root canal* AND (therapy OR procedure OR treatment OR retreatment OR management OR surgery OR filling OR debridement OR devitalization OR obturation OR preparation OR resection OR hemisection)) OR pulpectomy OR pulpotomy OR (pulp AND (revascularization OR regeneration OR capping)) OR apical surgery OR apicoectomy OR ((peri-radicular OR periradicular) AND surgery) OR Apexification OR Apexogenesis)

OVID (Cochrane Database of Systematic Reviews, Cochrane Central Register of Controlled Trials)

((pain OR pains OR painful* OR headache* OR cephalalg* OR cephalg* OR cephalodynia* OR glossodynia* OR glossalg* OR glossopyrosis OR toothache* OR tooth-ache* OR odontalg* OR neuralg* OR neurodynia OR earache* OR ear-ache* OR otalg* OR hypersens* OR hyperalg* OR allodyn* OR tooth sensitivity OR teeth sensitivity OR dental sensitivity OR dentin sensitivity OR ache OR aches OR aching OR soreness OR sore OR flare-up* OR flareup*) OR (exp Pain/ OR exp Somatosensory Disorders/ OR exp Neuralgia/)) AND ((psych* OR emotion* OR emotive OR cognition* OR cognitive* OR behavior* OR behaviour* OR mental* OR motivation* OR expectanc* OR expectation* OR anticipat* OR memory OR memories OR attitude* OR belief* OR appraisal* OR uncertaint* OR predictability OR controllability OR helplessness OR self-efficacy OR perceived control OR felt control OR locus of control OR loss of control OR positive experience* OR negative experience* OR threat* OR feeling* OR mood* OR fear* OR anxiet* OR anxious* OR hypervigilant* OR depression OR depressive* OR guilt* OR shame* OR embarrassment* OR distress* OR stress* OR worry OR worrisome OR disgust* OR negative affect* OR positive affect* OR affective state* OR phobi* OR odontophobia OR panic OR concern OR concerns OR avoidance OR reward* OR reinforc* OR vicarious* OR coping* OR learning* OR adaptation* OR optimism OR pessimism OR somatization OR somatisation OR somatoform OR neuroticism OR catastrophiz* OR catastrophis* OR nervous* OR personalit* OR mindful*) OR (exp Behavioral Disciplines and Activities/)) AND ((Endodontic* OR (root canal* AND (therapy OR procedure OR treatment OR retreatment OR management OR surgery OR filling OR debridement OR devitalization OR obturation OR preparation OR resection OR hemisection)) OR pulpectomy OR pulpotomy OR (pulp AND (revascularization OR regeneration OR capping)) OR apical surgery OR apicoectomy OR ((peri-radicular OR periradicular) AND surgery) OR Apexification OR Apexogenesis) OR (exp Endodontics/))

CINAHL

(pain OR pains OR "painful* OR headache*" OR cephalalg* OR cephalg* OR cephalodynia* OR glossodynia* OR glossalg* OR glossopyrosis OR toothache* OR tooth-ache* OR odontalg* OR neuralg* OR neurodynia OR earache* OR ear-ache* OR otalg* OR hypersens* OR hyperalg* OR allodyn* OR "tooth sensitivity" OR "teeth sensitivity" OR "dental sensitivity" OR "dentin sensitivity" OR ache OR aches OR aching OR soreness OR sore OR flare-up* OR flareup* OR (MH Pain+) OR (MH "Pain Measurement+") OR (MH Hyperalgesia+) OR (MH Allodynia+) OR (MH Hyperesthesia+) OR (MH Hypesthesia+) OR (MH Paresthesia+) OR (MH "Dentin Sensitivity+")) AND (psych* OR emotion* OR emotive OR cognition* OR cognitive* OR behavior* OR behaviour* OR mental* OR motivation* OR expectanc* OR expectation* OR anticipat* OR memory OR memories OR attitude* OR belief* OR appraisal* OR uncertaint* OR predictability OR controllability OR helplessness OR "self efficacy" OR self-efficacy OR "perceived control" OR "felt control" OR "locus of control" OR "loss of control" OR "positive experience*" OR "negative experience*" OR threat* OR feeling* OR mood* OR fear* OR anxiet* OR anxious* OR hypervigilant* OR depression OR depressive* OR guilt* OR shame* OR embarrassment* OR distress* OR stress* OR worry OR worrisome OR disgust* OR "negative affect*" OR "positive affect*" OR "affective state*" OR phobi* OR odontophobia OR panic OR concern OR concerns OR avoidance OR reward* OR reinforc* OR vicarious* OR coping* OR learning* OR adaptation* OR optimism OR pessimism OR somatization OR somatisation OR somatoform OR neuroticism OR catastrophiz* OR catastrophis* OR nervous* OR personalit* OR mindful* OR (MH Behavioral Sciences+) OR (MH "Health Belief Model+")) AND (Endodontic* OR (("root canal*") AND (therapy OR procedure OR treatment OR retreatment OR management OR surgery OR filling OR debridement OR devitalization OR obturation OR preparation OR resection OR hemisection)) OR pulpectomy OR pulpotomy OR (pulp AND (revascularization OR regeneration OR capping)) OR "apical surgery" OR apicoectomy OR ((peri-radicular OR periradicular) AND surgery) OR Apexification OR Apexogenesis OR (MH Endodontics+))

Web of Science - All Databases + Conference Proceedings Citation Index

TS=((pain OR pains OR painful* OR headache* OR cephalalg* OR cephalg* OR cephalodynia* OR glossodynia* OR glossalg* OR glossopyrosis OR toothache* OR tooth-ache* OR odontalg* OR neuralg* OR neurodynia OR earache* OR ear-ache* OR otalg* OR hypersens* OR hyperalg* OR allodyn* OR "tooth sensitivity" OR "teeth sensitivity" OR "dental sensitivity" OR "dentin sensitivity" OR ache OR aches OR aching OR soreness OR sore OR flare-up* OR flareup*) AND (psych* OR emotion* OR emotive OR cognition* OR cognitive* OR behavior* OR behaviour* OR mental* OR motivation* OR expectanc* OR expectation* OR anticipat* OR memory OR memories OR attitude* OR belief* OR appraisal* OR uncertaint* OR predictability OR controllability OR helplessness OR self-efficacy OR "perceived control" OR "felt control" OR "locus of control" OR "loss of control" OR "positive experience*" OR "negative experience*" OR threat* OR feeling* OR mood* OR fear* OR anxiet* OR anxious* OR hypervigilant* OR depression OR depressive* OR guilt* OR shame* OR embarrassment* OR distress* OR stress* OR worry OR worrisome OR disgust* OR "negative affect*" OR "positive affect*" OR "affective state*" OR phobi* OR odontophobia OR panic OR concern OR concerns OR avoidance OR reward* OR reinforc* OR vicarious* OR coping* OR learning* OR adaptation* OR optimism OR pessimism OR somatization OR somatisation OR somatoform OR neuroticism OR catastrophiz* OR catastrophis* OR nervous* OR personalit* OR mindful*) AND (Endodontic* OR (("root canal*") AND (therapy OR procedure OR treatment OR retreatment OR management OR surgery OR filling OR debridement OR devitalization OR obturation OR preparation OR resection OR hemisection)) OR pulpectomy OR pulpotomy OR (pulp AND (revascularization OR regeneration OR capping)) OR "apical surgery" OR apicoectomy OR ((peri-radicular OR periradicular) AND surgery) OR Apexification OR Apexogenesis))

Scopus – only in TITLE-ABS-KEY + Conference

(pain OR pains OR painful* OR headache* OR cephalalg* OR cephalg* OR cephalodynia* OR glossodynia* OR glossalg* OR glossopyrosis OR toothache* OR tooth-ache* OR odontalg* OR neuralg* OR neurodynia OR earache* OR ear-ache* OR otalg* OR hypersens* OR hyperalg* OR allodyn* OR "tooth sensitivity" OR "teeth sensitivity" OR "dental sensitivity" OR "dentin sensitivity" OR ache OR aches OR aching OR soreness OR sore OR flare-up* OR flareup*) AND (psych* OR emotion* OR emotive OR cognition* OR cognitive* OR behavior* OR behaviour* OR mental* OR motivation* OR expectanc* OR expectation* OR anticipat* OR memory OR memories OR attitude* OR belief* OR appraisal* OR uncertaint* OR predictability OR controllability OR helplessness OR "self efficacy" OR self-efficacy OR "perceived control" OR "felt control" OR "locus of control" OR "loss of control" OR "positive experience*" OR "negative experience*" OR threat* OR feeling* OR mood* OR fear* OR anxiet* OR anxious* OR hypervigilant* OR depression OR depressive* OR guilt* OR shame* OR embarrassment* OR distress* OR stress* OR worry OR worrisome OR disgust* OR "negative affect*" OR "positive affect*" OR "affective state*" OR phobi* OR odontophobia OR panic OR concern OR concerns OR avoidance OR reward* OR reinforc* OR vicarious* OR coping* OR learning* OR adaptation* OR optimism OR pessimism OR somatization OR somatisation OR somatoform OR neuroticism OR catastrophiz* OR catastrophis* OR nervous* OR personalit* OR mindful*) AND (Endodontic* OR (("root canal*") AND (therapy OR procedure OR treatment OR retreatment OR management OR surgery OR filling OR debridement OR devitalization OR obturation OR preparation OR resection OR hemisection)) OR pulpectomy OR pulpotomy OR (pulp AND (revascularization OR regeneration OR capping)) OR "apical surgery" OR apicoectomy OR ((peri-radicular OR periradicular) AND surgery) OR Apexification OR Apexogenesis)

ProQuest,

noft((pain OR pains OR painful* OR headache* OR cephalalg* OR cephalg* OR cephalodynia* OR glossodynia* OR glossalg* OR glossopyrosis OR toothache* OR tooth-ache* OR odontalg* OR neuralg* OR neurodynia OR earache* OR ear-ache* OR otalg* OR hypersens* OR hyperalg* OR allodyn* OR "tooth sensitivity" OR "teeth sensitivity" OR "dental sensitivity" OR "dentin sensitivity" OR ache OR aches OR aching OR soreness OR sore OR flare-up* OR flareup*) AND (psych* OR emotion* OR emotive OR cognition* OR cognitive* OR behavior* OR behaviour* OR mental* OR motivation* OR expectanc* OR expectation* OR anticipat* OR memory OR memories OR attitude* OR belief* OR appraisal* OR uncertaint* OR predictability OR controllability OR helplessness OR self-efficacy OR perceived-control OR felt-control OR "locus of control" OR "loss of control" OR "positive experience*" OR "negative experience*" OR threat* OR feeling* OR mood* OR fear* OR anxiet* OR anxious* OR hypervigilant* OR depression OR depressive* OR guilt* OR shame* OR embarrassment* OR distress* OR stress* OR worry OR worrisome OR disgust* OR "negative affect*" OR "positive affect*" OR "affective state*" OR phobi* OR odontophobia OR panic OR concern OR concerns OR avoidance OR reward* OR reinforc* OR vicarious* OR coping* OR learning* OR adaptation* OR optimism OR pessimism OR somatization OR somatisation OR somatoform OR neuroticism OR catastrophiz* OR catastrophis* OR nervous* OR personalit* OR mindful*) AND (Endodontic* OR (("root canal*") AND (therapy OR procedure OR treatment OR retreatment OR management OR surgery OR filling OR debridement OR devitalization OR obturation OR preparation OR resection OR hemisection)) OR pulpectomy OR pulpotomy OR (pulp AND (revascularization OR regeneration OR capping)) OR "apical surgery" OR apicoectomy OR ((peri-radicular OR periradicular) AND surgery) OR Apexification OR Apexogenesis))
