## Appendix 2 for "Psychological Contributors to Pain Before, During, and After Endodontic Procedures: A Scoping Review"

**Scales used for measurement of psychological variables:**

**Anxiety measurement for Pre-procedural pain:**

1. State Anxiety: Measured using State-Trait Anxiety Inventory (STAI).

2. Dental Anxiety: Measured in five studies using different scales, including:

- NRS (Numeric Rating Scale)

- DAS (Dental Anxiety Scale)

**Anxiety measurement scales related to procedural pain:**

1. Anxiety:

- STAI (State-Trait Anxiety Inventory)

- NRS (Numeric Rating Scale)

- CARS (Childhood Autism Rating Scale)

2. Dental Fear:

- DFS (Dental Fear Survey)

3. Dental Anxiety:

- DAS4 (Dental Anxiety Scale 4)

- MDAS (Modified Dental Anxiety Scale)

- SDAI (Spielberger Dental Anxiety Inventory)

- VAS (Visual Analog Scale)

- Questionnaires (Specific questionnaires designed to measure dental anxiety)

4. Discomfort and Stress:

- VAS (Visual Analog Scale)

**Anxiety scales related to post-procedural pain:**

1. Anxiety:

- State and Trait Anxiety:

- Numerical Rating Scale (NRS)

- HADS (Hospital Anxiety and Depression Scale)

2. Dental Anxiety:

- 11-point NRS (Numerical Rating Scale)

- Graded Chronic Pain Scale (GCPS)

- Questionnaire (Specific questionnaires designed to measure dental anxiety)

3. Pain Catastrophizing:

- Pain Catastrophizing Scale (PCS)

- Catastrophizing subscale of the Pain Coping Scale (PCS)

4. Oral Health-Related Anxiety:

- Oral Health Impact Profile Questionnaire (OHIP-14)

- PHQ-4, PHQ-15, and OHIP-49 (Specific questionnaires measuring discomfort, somatic symptoms, and psychological disability)

5. Stress:

- A question in a national survey (specific details about the question are not provided)

**Expected pain scales** **for procedural and post-procedural:**

1. Visual Analogue Scales (VAS): VAS was one of the tools used to measure expected pain.

2. Numerical Rating Scales (NRS): NRS was another tool used to assess expected pain.

3. Pain Expectation Scale (PES): The Pain Expectation Scale was used as a specific scale to measure pain expectation.

**Depression scales for procedural and post-procedural:**

1. Hospital Anxiety and Depression Scale (HADS): This scale was used in multiple studies to measure depression.

2. Beck Depression Inventory-II: This scale was used by (Pillai et al.) to assess depression.

3. Questionnaire: A questionnaire was used by Yang to measure depression.

**‘Positive treatment expectation’** **scales for procedural and post-procedural:**

1. Questionnaire: A questionnaire was used by (Daline et al.) to assess patients' optimism about the treatment outcome.

2. Numerical Rating Scale (NRS): NRS was used by (Wu et al.) to measure expected pain relief in patients undergoing emergency endodontic treatment.

**Personality in just procedural pain:**

"short form of the NEO Five-Factor Inventory scale."

**‘Desire for Control over Dental Treatment’ for just post-procedural pain:**

- Iowa Dental Control Index (ICDI): Used to assess the desire for control over dental treatment and felt control.

**Perception of Dentist for just postprocedural pain:**

- Baseline question to assess whether patients had ever had a painful, frightening, or embarrassing experience with a dentist: Used by (Maggiria and Locker).

- Assessment of the perceived empathy of the dentist during root canal treatment: Used by (Perkovic et al).

**‘Somatic Focus or Awareness’** **for just postprocedural pain:**

- Patient Health Questionnaire 15 (PHQ-15): Used by (Pillai et al.) to measure somatic symptom severity.

- Pennebaker Inventory of Limbic Languidness (PILL): Used by Applebaum and Maixner to measure somatization.

- National survey to measure melancholy, consultation with a psychiatrist, and suicidal thoughts: Used by (Yang et al).

**‘Pain Coping Strategies’ for just postprocedural pain:**

- Coping Strategies Questionnaire (CSQ): Used in two studies to measure pain coping strategies before emergency endodontic treatment.

**‘Beliefs about Pain’** **for just post-procedural pain:**

- Questionnaire assessing the relationship between "stress that makes the pain worse" and severe post-procedural pain: Used by Law.

**‘Positive and Negative Effects’** **for just postprocedural pain:**

- Profile of Mood States (POMS) questionnaire: Used in two studies to evaluate positive and negative affect related to dental treatment prior to emergency endodontic treatment.
